# Investigating the implications of COVID-19 for the rural and remote population of Northern Ontario using a mathematical model

**DOI:** 10.1101/2020.09.17.20196949

**Authors:** DW Savage, A Fisher, S Choudhury, R Ohle, RP Strasser, A Orkin, V Mago

**Author notes:** Corresponding Author: David Savage. Funding Statement: This study was partially funded by the Lakehead University VPRI Strategic Research Fund (Romeo #1467916) and the Northern Ontario Academic Medical Association (NOAMA) Clinical Innovation Opportunities Fund (C-14-2-18). Competing Interests: None. Author Contributions: DWS, AF, SC, and VM worked on all aspects of this study including the conceptualization, analysis, interpretation of data, drafting and revision of the manuscript and have approved the final submission. RO, RPS and AO provided interpretation, revisions to the manuscript and have approved the final submissions. All authors agree to be accountable for all aspects of the work.

## Abstract

**Background:** COVID-19 has the potential to disproportionately affect the rural, remote, and Indigenous populations who typically have a worse health status and live in substandard housing, often with overcrowding. Our aim is to investigate the potential effect of COVID-19 on intensive care unit (ICU) resources and mortality in northwestern Ontario.

**Methods:** This study was conducted in northwestern Ontario which has a population of 230,000. A set of differential equations were used to represent a modified Susceptible-Infectious-Recovered (SIR) model with urban and rural hospital resources (i.e., ICU and hospital beds). Rural patients requiring ICU care flowed into the urban ICU. Sensitivity analyses were used to investigate the effect of poorer health status (i.e., increased hospital admission, ICU admission, and mortality) and overcrowding (i.e., increased contact rate) in the rural population as compared to the urban population. Physical distancing within the urban population was modelled as a decreased contact rate.

**Results:** At the highest contact rate, the peak in daily active cases, ICU bed requirements and mortality was higher and occurred earlier than lower contact rates. The urban population with a lower contact rate and baseline health status had a lower predicted prevalence of active cases and lower mortality than the rural population.

**Interpretation:** An increased contact rate and worse health status in the rural population will likely increase the required ICU resources and mortality as compared to the urban population. Rural populations will likely be affected disproportionately more than urban populations.

## Introduction

The ongoing worldwide outbreak of the respiratory disease known as the novel coronavirus (COVID-19) has caused widespread mortality and morbidity leaving few countries unaffected. Countries and regions with populations that have a lower health status and/or fewer healthcare resources are at higher risk for the effects of COVID-19 (1). This disease appears to affect older adults and people with medical comorbidities disproportionately, however, younger adults and children are not spared (2).

Within Canada, people who live in rural and remote areas, including Indigenous populations have a substantial burden of illness and an overall lower health status in comparison with urban Canadians. Rural populations have a higher burden of diabetes, circulatory, and respiratory diseases and experience a 33% higher all-cause mortality compared to urban populations (3). Overall, these conditions contribute to lower life expectancy for this population (4). In addition, healthcare resources including equipment, infrastructure and personnel in rural areas is often limited (5,6). Although the vulnerability of rural and remote populations to COVID-19 has been identified conceptually, little work has been done to quantify or model the potential disease burden on this population(7).

Several modelling studies have been published examining the spread COVID-19 and hospital resources required for patients (8,9). These studies have shown significant spread of the disease, a lack of hospital capacity (e.g., hospital ward beds and ICU beds), and the benefit of interventions to decrease spread of the disease (e.g., physical distancing). In Ontario, Canada, the rate of spread of COVID-19 appears to be declining and the most recent projections for hospital and ICU capacity are more optimistic. However, given the high risk for recurrence of this disease, it is important to understand the potential risk for the remote, rural and Indigenous populations(10,11).

Much of the focus in Canada from a planning and policy perspective has been on projecting outcomes for urban areas with less consideration for rural populations. Northwestern Ontario, a large area with urban, rural, remote and Indigenous communities has significant variation in the social, economic and health status among its population. The aim of this study was 1) to assess, project and identify the risk of COVID-19 transmission, resource use and mortality in urban and rural communities in northwestern Ontario and 2) develop an online tool for policy planners and decision makers to use (https://covid.datalab.science). This study and the online tool will provide support to decision makers to reduce health inequity and respond proactively to the anticipated outcome disparities between urban and rural populations during this pandemic.

## Methods

This study used a system dynamics modelling framework to evaluate the effect of COVID-19 spread on urban and rural populations and the potential effect on hospital resources. All data and parameters used in the analysis were from publicly available sources; no research ethics board approval was required.

### Setting

The study was conducted in northwestern Ontario, a region 526,000 km^2^ in size. The population is 231,000 with 48% living in Thunder Bay, the only large urban centre (Figure 1). The region has 13 rural communities with 10 hospitals. For this study, the population in Thunder Bay will be referred to as the urban population and the remainder of the region will be referred to as the rural population. Most of the rural hospitals provide emergency and inpatient care delivered by rural generalist family physicians. Several of the larger rural communities (i.e., Sioux Lookout, Dryden, and Fort Frances) also provide some surgical and other specialist medicine services (e.g., general internal medicine, psychiatry, general surgery, obstetrics), but do not have dedicated critical care inpatient capacity. The primary economic driver in the region is the natural resource industry and outdoor tourism. There are 88 Indigenous communities in the region with 27% of the population being Indigenous. Many of these communities are only accessible by air. Much of the primary care for these remote communities is provided by nursing stations and family physicians located in Sioux Lookout.

**Figure 1.**
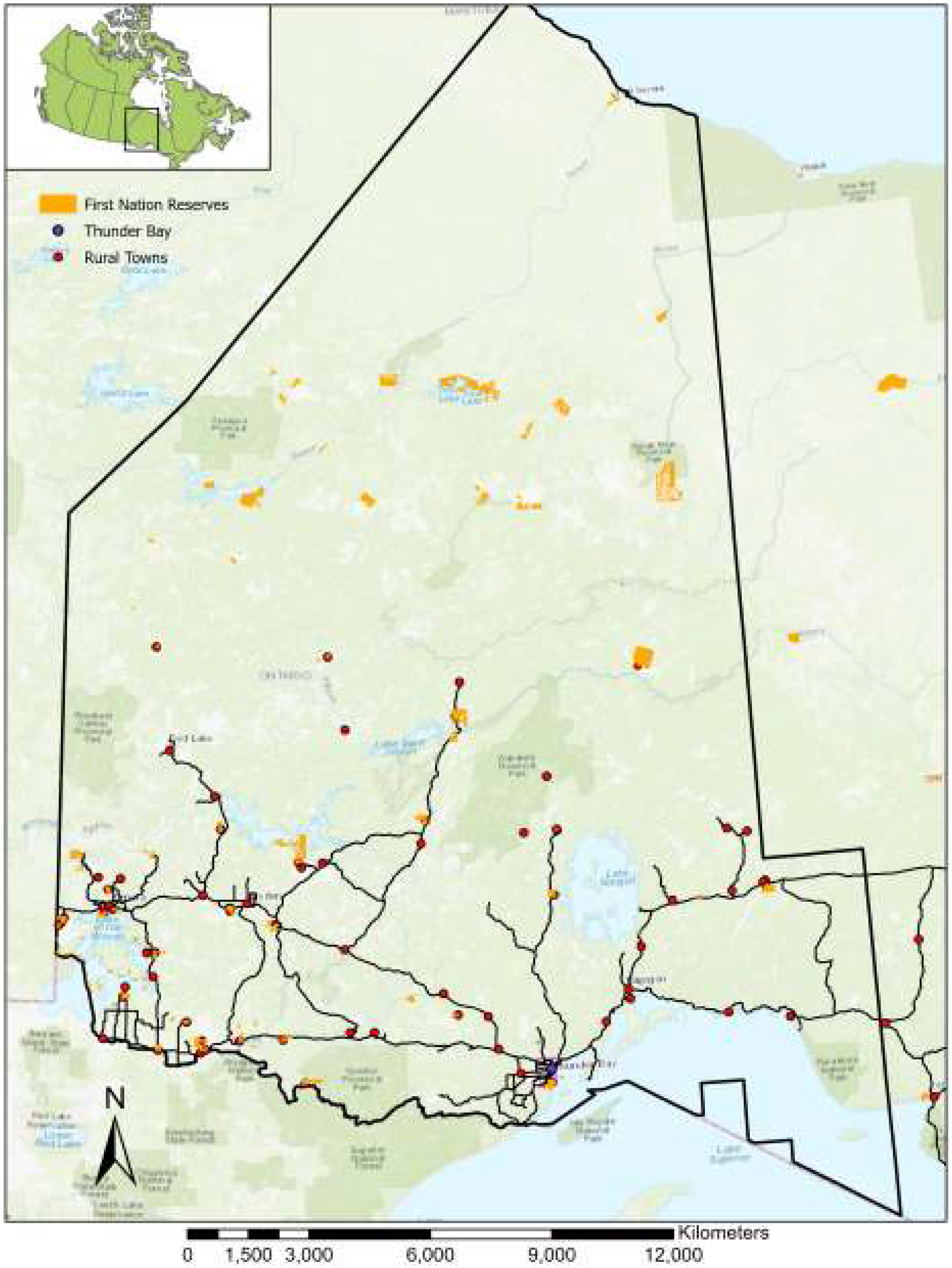
Map of northwestern Ontario showing the location of Thunder Bay, rural and First Nations communities.

The Thunder Bay Health Sciences Centre (TBRHSC) is the regional referral centre for northwestern Ontario and evaluates the majority of patients requiring acute or specialist care. A small proportion of patients from communities along the Manitoba border are assessed in Winnipeg. The TBRHSC is able to provide most of the medical and surgical needs for the region including a 22 bed Level 3 intensive care unit (ICU). During surges in patient demand, the ICU is able to accommodate 38 ventilated patients and 20 non-ventilated patients. Kenora, a city of approximately 15,000 does have a Level 2 ICU and is able to accommodate 8 ventilated patients during a surge in patient demand. The towns of Fort Frances, Dryden, and Sioux Lookout have access to approximately 11 ventilators in total and, with support from Thunder Bay, may be able to manage these patients during a surge in demand.

### Model

The tool developed was structured as a system dynamics model using differential equations to describe the infectious disease spread and healthcare resources available (See Appendix 1 for model description and equations). COVID-19 spread was modelled using a modified Susceptible-Infectious-Recovered (SIR) modelling framework with parameters describing the initial susceptible population and spread of the disease (Table 1). Hospital bed and ICU resources were connected to the spread model to show the potential pathway for a COVID-19 patient (Figure 2). To examine disease spread, healthcare policies and resources in the region, two sub-models were created to represent the rural population and another to represent the urban population. COVID-19 spread is modelled between the two sub-models. This allows different susceptible populations, contact rates, hospital and ICU admission rates and mortality rates that may vary between these populations. The urban sub-model has the majority of ICU resources and would accept all rural patients until capacity was met, at which point the available rural ICU beds would begin to fill. If all ICU beds are filled, a user defined percentage of patients will flow into a death state and the remainder will survive. Hospitalized patients not requiring ICU care will remain in their respective communities. Model parameters were obtained from published literature and the Canadian federal government COVID-19 projections (22).

**Table 1.** Modelling parameters and references.

| Parameter | Value | Description | Reference |
| --- | --- | --- | --- |
| Total population:<br>Rural<br>Urban | 120,000<br>110,000 | The total rural and urban population within northwestern Ontario. | (26,27) |
| Number of initial infected individuals:<br>Rural<br>Urban | 20<br>10 | The number of initial infected individuals within the population. | Estimated |
| Infectivity | 0.026 | Calculated from $R_0 = 2.2$(28), Contact rate = 12, and Infectious period = 7 days. | Estimated |
| Contact rate | 12 | Average number of interactions a person has per day. | (12) |
| Infectious Period | 7 days | Number of days the infection can spread from person to person. | (29) |
| ICU capacity (total beds):<br>Rural<br>Urban | 17<br>40 | Northwestern Ontario has 4 rural and 22 urban ICU beds with additional capacity added for potential patient surges. |  |
| Population susceptibility rate | 10% | Percentage of the population susceptible to COVID-19. | Estimated |
| Hospital admission rate | 8% | The percentage of patients infected that will be admitted to hospital. | (22) |
| ICU admission rate | 32% | The percentage of patients admitted to hospital that will require ICU level care. | (22) |
| Hospital length of stay | 10 days | Number of days a patient will be admitted to the hospital ward bed. | (30) |
| ICU length of stay | 21 days | Number of days a patient will be admitted to the ICU bed. | (31) |
| ICU mortality rate | 48% | The mortality rate for the patients admitted to the ICU. | (22) |
| ICU overflow mortality rate | 80% | If the ICU reached capacity and patients were unable to receive care, a specified percentage of patients move to the death state with the remainder surviving in the hospital ward. | Estimated |
| Post-ICU discharge length of stay in hospital | 7 days | Length of stay for those patients transferred from the ICU to the hospital ward. | Estimated |

**Figure 2.**
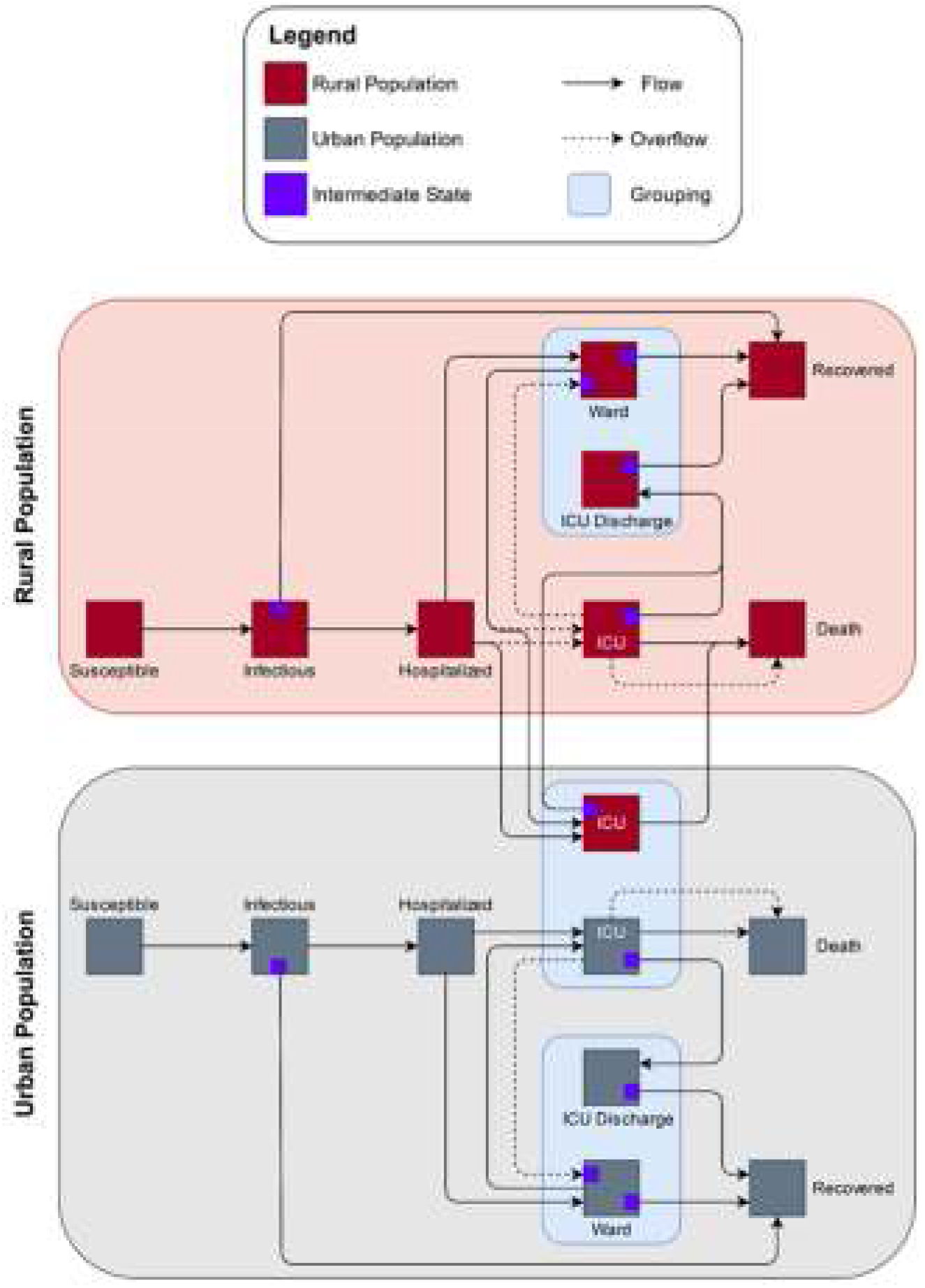
COVID-19 model structure for the rural and urban populations of northwestern Ontario. All potential patients start in the susceptible state and finish in either the recovered or death state. Disease spread (i.e., contact rate) was modelled independently between populations. Patients requiring hospital admission, ICU admission and mortality rate were also modelled independently between the two populations. Rural patients requiring ICU care initially flow into the urban ICU. However, if the urban ICU reaches capacity then the overflow policy has rural patients remaining in the rural communities. If all ICUs reach capacity, another overflow policy specifies that a proportion of patients are transferred to the death state with the remaining patients surviving in the ward. ICU and ward groupings were used within the model to track patients transferred between the rural and urban populations to ensure accurate measurement of ICU bed requirements and mortality between the populations. An intermediate state was used in the model for states (e.g., ward, ICU, etc.) where the patient must remain in the state for a specified length of time.

### Modelling rural overcrowding and health status

Using our model, we completed sensitivity analyses to identify the effect of poorer health status (i.e., increased hospital admission, ICU admission, and mortality) and overcrowding (i.e., increased contact rate) in the rural population as compared to the urban population. Health status in the model was represented by varying the hospital admission, ICU admission and mortality rates. In the urban population, the base parameters from the literature were used to represent their better health status. For the rural population, we incrementally increased the rates of hospital admission, ICU admission and mortality by 10%, 20% and 30% to model the poorer health status (Table 2).

**Table 2.**
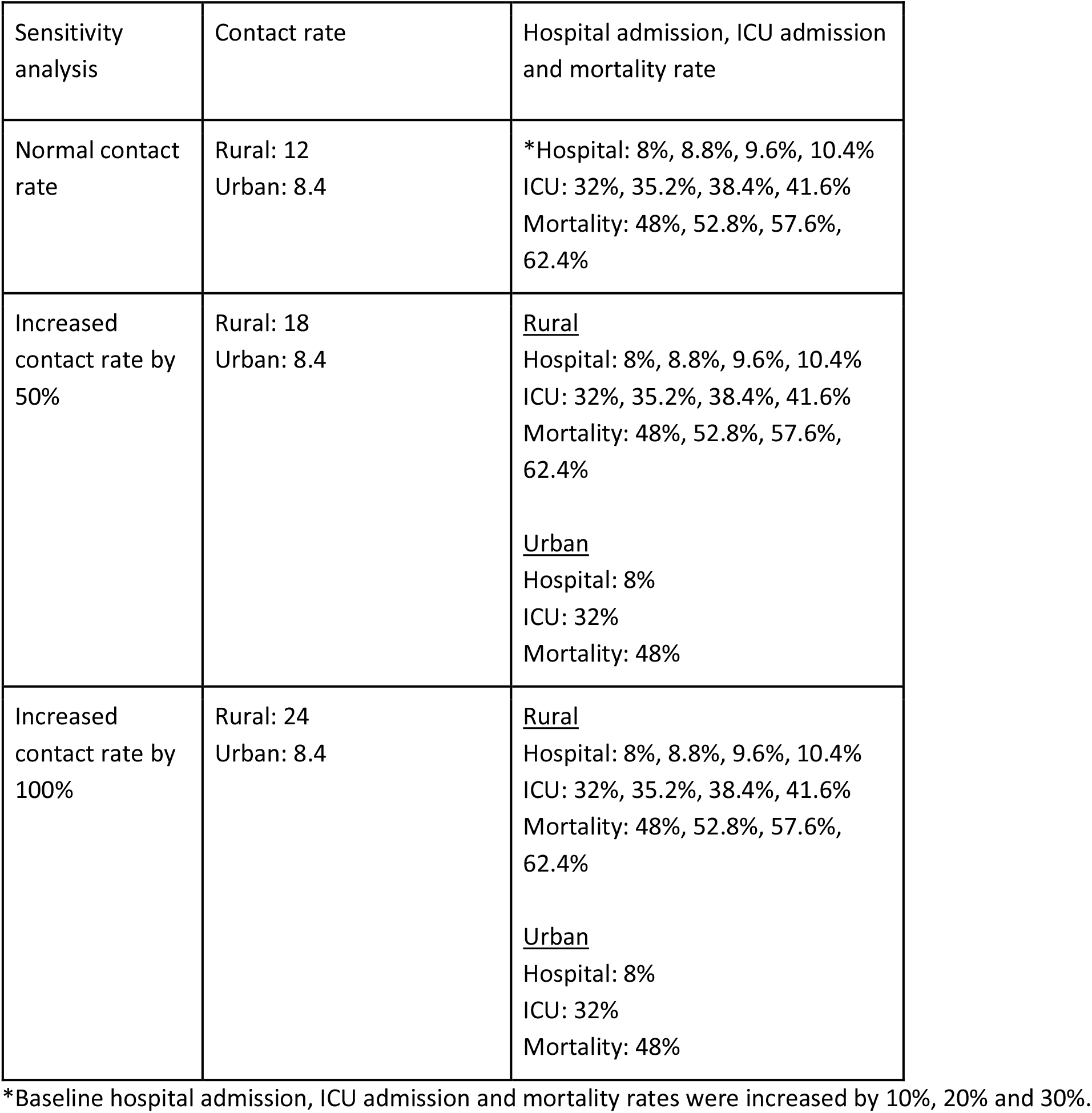
Summary of parameters used in the sensitivity analyses to examine effect of COVID-19 on urban and rural populations.

| Sensitivity analysis | Contact rate | Hospital admission, ICU admission and mortality rate |
| --- | --- | --- |
| Normal contact rate | Rural: 12<br>Urban: 8.4 | *Hospital: 8%, 8.8%, 9.6%, 10.4%<br>ICU: 32%, 35.2%, 38.4%, 41.6%<br>Mortality: 48%, 52.8%, 57.6%, 62.4% |
| Increased contact rate by 50% | Rural: 18<br>Urban: 8.4 | <u>Rural</u><br>Hospital: 8%, 8.8%, 9.6%, 10.4%<br>ICU: 32%, 35.2%, 38.4%, 41.6%<br>Mortality: 48%, 52.8%, 57.6%, 62.4%<br><u>Urban</u><br>Hospital: 8%<br>ICU: 32%<br>Mortality: 48% |
| Increased contact rate by 100% | Rural: 24<br>Urban: 8.4 | <u>Rural</u><br>Hospital: 8%, 8.8%, 9.6%, 10.4%<br>ICU: 32%, 35.2%, 38.4%, 41.6%<br>Mortality: 48%, 52.8%, 57.6%, 62.4%<br><u>Urban</u><br>Hospital: 8%<br>ICU: 32%<br>Mortality: 48% |

\*Baseline hospital admission, ICU admission and mortality rates were increased by 10%, 20% and 30%.

The average number of social contacts per day for this study was estimated to be 12 (12). The overcrowding in the rural population that is observed in some homes (i.e., especially in First Nations) was assessed by increasing the contact rate by 50% and 100%. The physical distancing observed in the urban population was modelled by decreasing the contact rate by 30%. COVID-19 spread was modelled over a 120 day period to demonstrate the predicted outbreak pattern and resources required. Given the study’s focus on overcrowding and rural health status, the outcomes of interest are the infection rate, ICU admission rate and mortality rate.

## Results

The results from each model run are available online for preview (https://datalab-covid.ddns.net:9001/login; Password: rYwaLoAt). The number of active cases that we modelled in the rural population more than doubled (i.e., approximately 250 to 500 active cases per 10,000) when the contact rate was increased from 12 to 24 and also peaked in frequency sooner (i.e., day 17 vs. day 40) (Figure 3). The urban population with physical distancing modelled with a contact rate of 8.4 showed fewer active cases and a longer infectious period. Since the health status proxy we use does not affect the disease spread, there was little variation in the number of active cases (i.e., the range between the upper and lower bound was small).

**Figure 3.**
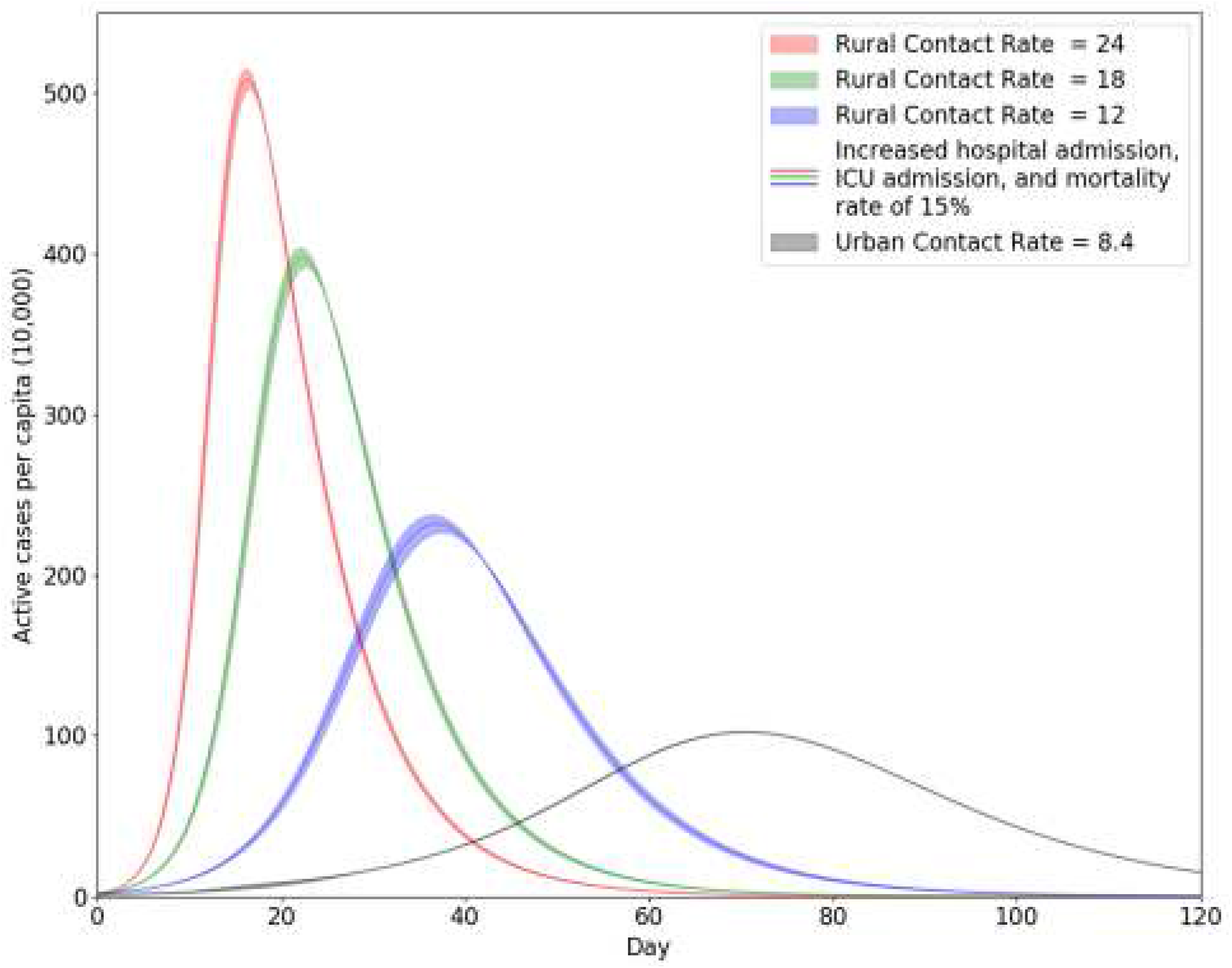
The projected number of active cases of COVID-19 in the rural and urban population. The three levels of contact rate for the rural population are 12, 18, and 24. The shaded regions demonstrate the range of health statuses examined with the lower bound corresponding to the baseline hospital admission, intensive care unit (ICU) admission and mortality rate while the upper bound represents a 30% increase in these rates.

The highest contact rate of 24 showed the earliest and highest peak in the number of ICU beds required (i.e., range of 108 to 130) while the average contact rate of 12 peaked later and showed a range of required ICU beds from 72 to 84 (Figure 4). The observed range in ICU beds required for a particular contact rate resulted from the range of health statuses examined. For all three contact rates investigated, the number of required ICU beds was influenced by the late rise in the number of infections in the urban population on approximately day 40.

**Figure 4.**
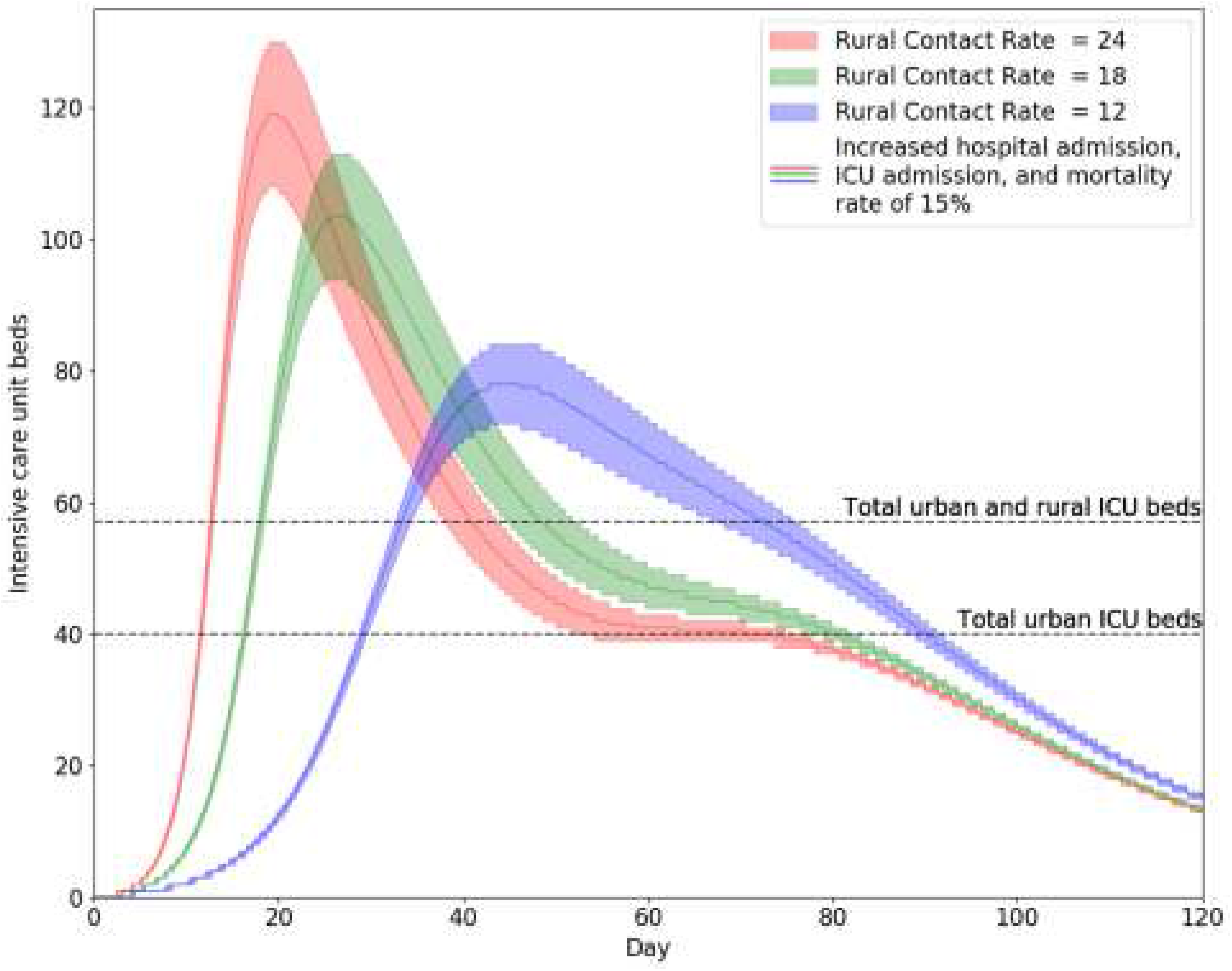
The total number of ICU beds needed for the urban and rural population combined. The three levels of contact rate for the rural population are 12, 18, and 24. The shaded regions demonstrate the range of health statuses examined with the lower bound corresponding to the baseline hospital admission, intensive care unit (ICU) admission and mortality rate while the upper bound represents a 30% increase in these rates. The total number of available urban ICU beds is noted along with the total number of ICU beds in the region.

The mortality projection in this study demonstrated a similar trend to the number of active cases and the number of required ICU beds (Figure 5). With a contact rate of 24 in the rural population, the mortality peaked on day 14 with a range of 1.9 to 3.4 deaths per 10,000 based on the range of health statuses examined. The lowest contact rate of 12 in the rural population predicted a mortality range of 0.6 to 1.0 deaths per 10,000. In the urban population, the mortality was 0.2 deaths per 10,000 in the population.

**Figure 5.**
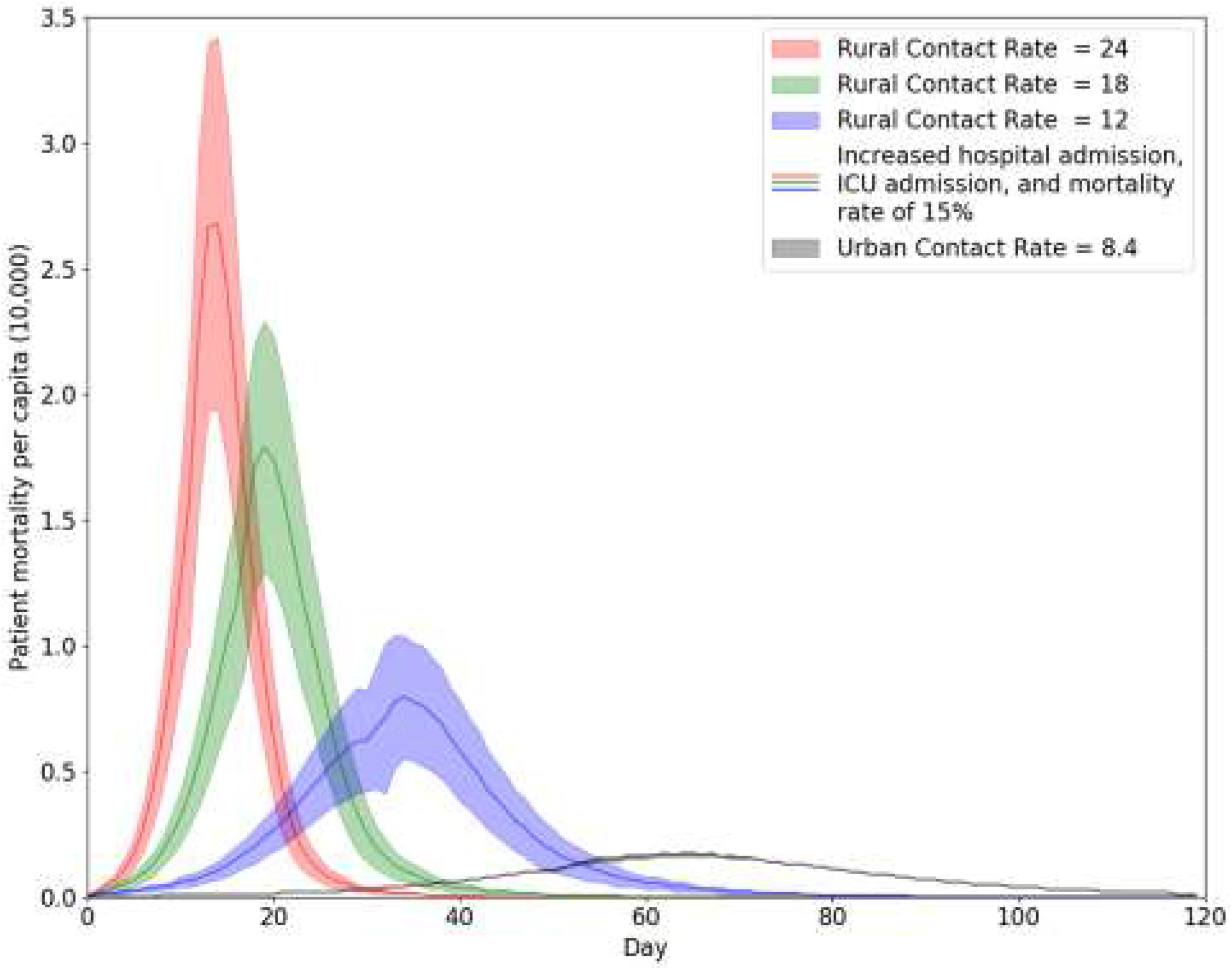
The patient mortality due to COVID-19 in the rural and urban population. The three levels of contact rate for the rural population are 12, 18, and 24. The shaded regions demonstrate the range of health statuses examined with the lower bound corresponding to the baseline hospital admission, intensive care unit (ICU) admission and mortality rate while the upper bound represents a 30% increase in these rates.

### Interpretation

This study provides a comparison of predicted urban and rural outcomes for COVID-19 infections, ICU admissions and mortality rate, and highlights the potential disproportionate effect this disease may have on people living in rural, remote and Indigenous communities. Given the current disease spread rates in northwestern Ontario, we will have sufficient ventilator capacity to meet demands for the first wave of disease transmission. However, the expected recurrence of this disease, potentially during the Influenza season may cause a more severe outbreak with a serious effect on the rural populations.

### Health of Rural Populations

Much of the increased risk of COVID-19 for the rural population is related to lower social, economic, and health status within these populations. These populations have higher burdens of chronic illness and a higher proportion of individuals over 65 years of age. From a COVID-19 perspective, epidemiological data already shows that older patients with chronic illness are at increased mortality risk (13). In the United States, some marginalized populations share many of the same social, economic and health challenges of Canada’s rural populations and have already been disproportionately affected by this disease from both an infection and mortality rate (14). Health status in this study was incorporated by increasing the rates of hospital admission, ICU admission and mortality. The precise effect of poorer health in the rural population is difficult to quantify, however, by conducting a sensitivity analysis we can generate a range of possible scenarios. These scenarios can then be used to test different policy options that potentially improve outcomes for rural populations.

The central public health intervention during this pandemic has been physical distancing to reduce contact rate amongst the population and thus disease spread(12). However, in many rural areas, especially remote First Nations, housing shortages have caused overcrowding with an increase in the average number of people living in a single dwelling (15). Overcrowding in homes has been linked to illnesses such as Tuberculosis in Indigenous adults and lower respiratory tract infections in children (16,17). In addition to housing challenges, many remote First Nation communities also experience food insecurity and a lack of potable water(18).

### Healthcare Resources

Prior to the global spread of COVID-19 cases and the ICU surge planning for Ontario, the northwest region had 11.3 ventilators per 100,000 in the population as compared to the Ontario average of 12.4 (19). In addition to fewer ventilators per capita, the people living in rural areas must travel long distances by either ground or air ambulance to access care. The Indigenous peoples living on northern First Nations (i.e., approximately 30,000 people) often only have access to a nursing station and must board a plane to see a physician in Sioux Lookout(20) (21). The modelling approach and parameterization in this study addresses this disparity in healthcare resources for the rural population and shows that they will likely have a higher mortality than the urban population. Current projections of COVID-19 spread and mortality favour more optimistic outcomes as compared to some of the earlier projections(22).

### Healthcare Policy

The rural, remote and Indigenous populations of northwestern Ontario are at high risk for increased morbidity and mortality from COVID-19. Much of the modelling and projections published ignore the large disparity in health status and available healthcare resources for the rural populations. Policy makers must acknowledge that these populations could potentially have much worse outcomes than urban populations. They share many of the same challenges as populations in the US that are marginalized and have significant health disparities (14,23). Rural, remote, and Indigenous communities require targeted policies for disease monitoring and interventions that not only recognize their unique challenges in terms of social, economic, and health status but also the role of geography in their outcomes(24). Potential targeted policies may include greater local testing, tracing, monitoring and treatment capacity as well as early planning for enhanced ventilator capacity in small rural hospitals, telemedicine and transport planning for regional management of surges in ventilator demand.

### Future work

Future research investigating the effect of COVID-19 on rural, remote and Indigenous communities will be important. Developing models and planning tools for allocating resources including personnel, equipment, and infrastructure will be important if outbreaks occur throughout Canada at different times and resources need to be shifted to meet demand.

### Study Limitations

The number of predictions of COVID-19 spread, morbidity and mortality from modelling studies has grown monthly. Unfortunately, parameterization of these models can be difficult. Epidemiological research into COVID-19 is still in the early stages given that this disease has spread quickly throughout the world. Our understanding of the variation and severity of symptoms is limited which makes the estimation of the worldwide infection rate difficult. Many patients will have mild symptoms and will be self-isolating based on the public health policies of various countries(25). These patients may not be accounted for in the estimation of hospital admission, ICU admission, and mortality rates and therefore these rates may actually be significantly lower. As these and many other parameters are better estimated in the coming months, updates to modelling projections should be made.

## Data Availability

All data is available in the paper or the published sources.

https://covid.datalab.science/

## Acknowledgements

This study was partially funded by the Lakehead University VPRI Strategic Research Fund (Romeo #1467916) and the Northern Ontario Academic Medical Association (NOAMA) Clinical Innovation Opportunities Fund (C-14-2-18). Thanks to Alex Bilyk and LU-CARIS for producing the map showing the study area.

## References

1. Ji Y, Ma Z, Peppelenbosch MP, Pan Q. Potential association between COVID-19 mortality and health-care resource availability. Lancet Glob Health. 2020 Apr;8(4):e480.

2. Wu Z, McGoogan JM. Characteristics of and important lessons from the coronavirus disease 2019 (COVID-19) outbreak in China: summary of a report of 72 314 cases from the Chinese Center for Disease Control and Prevention. JAMA [Internet]. 2020; Available from: https://jamanetwork.com/journals/jama/article-abstract/2762130

3. CIHI. How Healthy Are Rural Canadians? An Assessment of Their Health Status and Health Determinants, A Component of the Initiative Canada Rural Communities: Understanding Rural Health and Its Determinants [Internet]. CIHI. 2006 [cited 2020 Apr 9]. Available from: https://secure.cihi.ca/free_products/rural_canadians_2006_report_e.pdf

4. Manuel DG, Goel V, Williams JI, Corey P. Health-adjusted life expectancy at the local level in ontario. Chronic Dis Can. 2000;21(2):73–80.

5. Law MR, Dijkstra A, Douillard JA, Morgan SG. Geographic accessibility of community pharmacies in ontario. Healthc Policy. 2011 Feb;6(3):36–46.

6. Hay C, Pacey M, Bains N, Ardal S. Understanding the unattached population in Ontario: evidence from the Primary Care Access Survey (PCAS). Healthc Policy. 2010;6(2):33.

7. Monnat S. Why coronavirus could hit rural areas harder. Issue Brief. 2020;16.

8. Tuite AR, Fisman DN, Greer AL. Mathematical modelling of COVID-19 transmission and mitigation strategies in the population of Ontario, Canada. CMAJ [Internet]. 2020 Apr 8; Available from: http://dx.doi.org/10.1503/cmaj.200476

9. Kucharski AJ, Russell TW, Diamond C, Liu Y, Edmunds J, Funk S, et al. Early dynamics of transmission and control of COVID-19: a mathematical modelling study. Lancet Infect Dis [Internet]. 2020 Mar 11; Available from: http://dx.doi.org/10.1016/S1473-3099(20)30144-4

10. Xu S, Li Y. Beware of the second wave of COVID-19. Lancet. thelancet.com; 2020 Apr 25;395(10233):1321–2.

11. Tuite A, Greer AL, De Keninck S, Fisman DN. Reduced COVID-19-Related Critical Illness and Death, and High Risk of Epidemic Resurgence, After Physical Distancing in Ontario, Canada. medRxiv [Internet]. 2020; Available from: https://www.medrxiv.org/content/10.1101/2020.04.29.20084475v1.abstract

12. Ferguson N, Laydon D, Nedjati Gilani G, Imai N, Ainslie K, Baguelin M, et al. Report 9: Impact of non-pharmaceutical interventions (NPIs) to reduce COVID19 mortality and healthcare demand. 2020; Available from: https://spiral.imperial.ac.uk/handle/10044/1/77482

13. Grasselli G, Zangrillo A, Zanella A, Antonelli M, Cabrini L, Castelli A, et al. Baseline Characteristics and Outcomes of 1591 Patients Infected With SARS-CoV-2 Admitted to ICUs of the Lombardy Region, Italy. JAMA [Internet]. 2020 Apr 6; Available from: http://dx.doi.org/10.1001/jama.2020.5394

14. Yancy CW. COVID-19 and African Americans. JAMA [Internet]. 2020 Apr 15; Available from: http://dx.doi.org/10.1001/jama.2020.6548

15. First Nations Information Governance Centre, National Report of the First Nations Regional Health Survey Phase 3: Volume Two, (Ottawa: 2018). 1 pages. Published in July 2018.

16. Clark M, Riben P, Nowgesic E. The association of housing density, isolation and tuberculosis in Canadian First Nations communities. Int J Epidemiol. 2002 Oct;31(5):940–5.

17. Banerji A, Panzov V, Young M, Robinson J, Lee B, Moraes T, et al. Hospital admissions for lower respiratory tract infections among infants in the Canadian Arctic: a cohort study. CMAJ Open. 2016 Oct;4(4):E615–22.

18. SLFNA. Sioux Lookout First Nations Health Authority: Our Children and Youth Health Report 2018 [Internet]. Sioux Lookout First Nations Health Authority [Internet]. Our Children and Youth Health Report 2018 [cited April 21, 2020]. Available from: 2018 [cited 2020 Apr 9]. Available from: https://slfnha.com/application/files/6215/4022/0883/CHSR_FINAL_-_Web_Version.pdf

19. Ontario Critical Care Lhin. Inventory of Critical Care Services: An Analysis of LHIN-Level Capacities [Internet]. Ministry of Health and Long-Term Care. 2006 [cited 2020 Apr 20]. Available from: https://www.criticalcareontario.ca/EN/Toolbox/Overview%20of%20Ontarios%20Critical%20Care%20System/Inventory%20of%20Critical%20Care%20Services%20%28An%20Analysis%20of%20LHIN-Level%20Capacities%29%20%282006%29.pdf

20. Matsumoto C-L, O’Driscoll T, Lawrance J, Jakubow A, Madden S, Kelly L. A 5 year retrospective study of emergency department use in Northwest Ontario: a measure of mental health and addictions needs. CJEM. 2017 Sep;19(5):381–5.

21. Terry O, Sharen Madden MD, Blakeloch B, Len Kelly MD, Others. Characterizing high-frequency emergency department users in a rural northwestern Ontario hospital: a 5-year analysis of volume, frequency and acuity of visits. Can J Rural Med. 2018;23(4):99–105.

22. Public health agency of Canada. COVID-19 in Canada: Modelling Update [Internet]. Aprile 28 2020 [cited 2020 Apr 28]. Available from: https://www.canada.ca/content/dam/phac-aspc/documents/services/diseases/2019-novel-coronavirus-infection/using-data-modelling-inform-eng-04-28.pdf

23. Tsai J, Wilson M. COVID-19: a potential public health problem for homeless populations. Lancet Public Health. 2020 Apr;5(4):e186–7.

24. Romanow R, Others. Building on Values: Report of the Commission on the Future of Health Care in Canada. 2002; Available from: https://qspace.library.queensu.ca/handle/1974/6882

25. Guan W-J, Ni Z-Y, Hu Y, Liang W-H, Ou C-Q, He J-X, et al. Clinical Characteristics of Coronavirus Disease 2019 in China. N Engl J Med. 2020 Apr 30;382(18):1708–20.

26. Can S. Census Profile, 2016 Census Northwest [Economic region], Ontario and Ontario [Province] [Internet]. Statistics Canada. 2017 [cited 2020 May 21]. Available from:https://www12.statcan.gc.ca/census-recensement/2016/dp-pd/prof/details/page.cfm?Lang=E&Geo1=ER&Code1=3595&Geo2=PR&Code2=35&SearchText=Northwest&SearchType=Begins&SearchPR=01&B1=All&GeoLevel=PR&GeoCode=3595&TABID=1&type=0

27. Can S. Census Profile, 2016 Census Thunder Bay [Census metropolitan area], Ontario and Ontario [Province] [Internet]. Statistics Canada. 2017 [cited 2020 May 21]. Available from: https://www12.statcan.gc.ca/census-recensement/2016/dp-pd/prof/details/page.cfm?Lang=E&Geo1=CMACA&Code1=595&Geo2=PR&Code2=35&Data=Count&SearchText=Thunder%20Bay&SearchType=Begins&SearchPR=01&B1=All

28. Liu Y, Gayle AA, Wilder-Smith A, Rocklöv J. The reproductive number of COVID-19 is higher compared to SARS coronavirus. J Travel Med [Internet]. 2020 Mar 13;27(2). Available from: http://dx.doi.org/10.1093/jtm/taaa021

29. Lin H, Liu W, Gao H, Nie J, Fan Q. Trends in Transmissibility of 2019 Novel Coronavirus-Infected Pneumonia in Wuhan and 29 Provinces in China [Internet]. 2020 [cited 2020 Apr 30]. Available from: https://papers.ssrn.com/abstract=3544821

30. Wang D, Hu B, Hu C, Zhu F, Liu X, Zhang J, et al. Clinical Characteristics of 138 Hospitalized Patients With 2019 Novel Coronavirus–Infected Pneumonia in Wuhan, China. JAMA. 2020 Mar 17;323(11):1061–9.

31. Yang X, Yu Y, Xu J, Shu H, Xia J’an, Liu H, et al. Clinical course and outcomes of critically ill patients with SARS-CoV-2 pneumonia in Wuhan, China: a single-centered, retrospective, observational study. Lancet Respir Med. 2020 May;8(5):475–81.

